# Investigating the Polygenic Relationship Between Cannabis Use and Schizophrenia in the All of Us Research Program

**DOI:** 10.1101/2025.05.20.25327979

**Authors:** Isabelle Austin-Zimmerman, Hayley HA Thorpe, John J Meredith, Jibran Khokhar, Tian Ge, Marta Di Forti, Arpana Agrawal, Emma C Johnson, Sandra Sanchez-Roige

## Abstract

**Objective:** Decades of research have identified a strong association between heavy cannabis use and schizophrenia, with evidence of correlated genetic factors. However, many studies on the genetic relationship between cannabis use and psychosis have lacked data on both phenotypes within the same individuals, creating challenges due to unmeasured confounding. We aimed to address this by using multi-modal data from the All of Us Research Program, which contains genetic data as well as information on schizophrenia diagnosis and cannabis use.

**Methods:** We tested the association between cannabis use disorder (**CUD**) and schizophrenia polygenic scores (**PGS**) and schizophrenia and heavy cannabis use. We tested models where both CUD and schizophrenia PGS were included as joint predictors of heavy cannabis use and schizophrenia case status. We defined three sets of cases based on comorbidities: relaxed (assessing for only the primary condition), strict (excluding for both conditions), and dual-comorbidity (including both conditions).

**Results:** CUD and schizophrenia polygenic liability were independently associated with heavy cannabis use; the schizophrenia PGS effect was very modest. In contrast, both schizophrenia and CUD PGS were independently associated with schizophrenia, with independent significant effects of CUD PGS. Polygenic liability to CUD was associated with schizophrenia in individuals without a documented history of cannabis use, suggesting widespread pleiotropy.

**Conclusions:** These findings underscore the need for comprehensive models that integrate genetic risk factors for heavy cannabis use to advance our understanding of schizophrenia aetiology.

## INTRODUCTION

Chronic cannabis use is associated with greater psychosis risk^1–4^, psychotic-like experiences^5–7^, and earlier age of onset for psychotic symptoms^5,6^. Many prior studies have argued that heavy cannabis use increases the risk of psychosis^1,2,4^.

A complementary hypothesis posits that common genetic mechanisms underlie cannabis use and psychosis^7–10^. Genome-wide association studies (**GWAS**) and twin studies support that schizophrenia, the most common psychotic disorder, is heritable^11,12^ as are lifetime cannabis use^13^ and cannabis use disorder (**CUD**)^14,15^. Cannabis use traits and schizophrenia are positively genetically correlated (*r_g_*range = 0.25-35^7^). The most recent Mendelian randomisation studies support a bidirectional causal relationship between schizophrenia and CUD^15,16^, with a greater magnitude of causal effect from CUD to schizophrenia. Further, with the advent of polygenic scores (**PGS**), which aggregate the effect sizes from GWAS across genome-wide variants to quantify genetic liability for a trait, it has been observed that schizophrenia (**SCZ**) PGS are associated with psychotic-like experiences in cannabis users users^17^. This indicates that genetic risk for schizophrenia might make one particularly vulnerable to the psychotogenic effects of cannabis use. Thus, delineating the genetic relationship between cannabis use and schizophrenia may inform psychosis prevention, particularly in the context of expanding cannabis legalization and accessibility.

Another complicating factor is that while the prevalence of cannabis use and CUD in individuals with schizophrenia is high^2–4,18–20^, no GWAS of schizophrenia account for this co-occurrence. Likewise, GWAS of CUD rarely account for comorbid psychosis unless such conditions are exclusionary criteria for study recruitment. Thus, it is plausible that the genetic correlation between schizophrenia and cannabis use/CUD is partially confounded by their co-occurrence in the population.

In the present study, we availed of a unique opportunity to disarticulate the schizophrenia - cannabis comorbidity by leveraging the multi-modal data of the National Institute of Health’s *All of Us* (**AoU**) Research Program, for which phenotypic data relevant to cannabis and schizophrenia and genetic data are available for up to ∼250,000 participants in the version 7 release. Using data from prior well-powered GWAS, we examined the association between CUD^15^ and schizophrenia (**SCZ**)^11,12^ PGS and cannabis-related traits and schizophrenia in the AoU cohort. We hypothesized that polygenic liability to both CUD and SCZ would exert independent effects on cannabis use and schizophrenia. However, we also expected that the associations would be attenuated in the absence of the comorbid condition (e.g., SCZ PGS would not strongly relate to cannabis-related traits without comorbid psychosis).

## METHODS

### Participants

Participants enrolled in AoU (v7) were included in our analysis. AoU is a diverse database of electronic health records (**EHR**), survey responses, physical measurements, and whole-genome sequencing data from blood or saliva samples; the v7 release included data for over ∼250,000 adults in the United States. For details on recruitment and genomic data^21–23^.

Unrelated participants who were assigned male or female at birth, had applicable EHR and/or survey-level data, had whole-genome sequencing (**WGS**) data, and whose genomes were statistically correlated with the genomes of European (**EUR**) or African (**AFR**) reference populations were included in the study. Genetic similarity was based on principal components (**PCs**) provided by AoU. Related participants were removed using a relatedness flagged sample list provided by AoU kinship estimation^21^.

Heavy cannabis use and schizophrenia were derived using multi-modal data including diagnostic codes (SNOMED Clinical Terms), survey data, and prescription medication records (**Table 1**). We defined heavy cannabis use as anyone with a diagnosis of cannabis use disorder as well as anyone who reported daily cannabis use within the last three months. We included daily cannabis use based on its strong correlations with CUD^13^ and with schizophrenia^3,4^. We defined schizophrenia as anyone with schizophrenia based on diagnostic codes and/or survey data. Participants were excluded from the control groups if they had a record of antipsychotic prescriptions. See **Supplementary Figure 1** for a participant inclusion flowchart.

**Table 1.** All of Us concepts and concept codes used to assign binary identifiers for schizophrenia, heavy cannabis use, heavy tobacco smoking, and antipsychotic medication statuses. Participants across different Concept IDs may overlap. See Supplementary Figure 1 for sample details.

| Phenotype | Concept ID | Source | N<sub>cases</sub> | Concept |
| --- | --- | --- | --- | --- |
| Schizophrenia | 435783 | Conditions | 2,445 | Schizophrenia |
|  | 43530520 | Personal and Family Health Survey | 440 | About how old were you when you were first told you had schizophrenia? |
|  | 43528847 | Personal and Family Health Survey | 335 | Are you currently prescribed medications and/or receiving treatment for schizophrenia? |
|  | 43530362 | Personal and Family Health Survey | 347 | Are you still seeing a doctor or health care provider for schizophrenia? |
| Heavy cannabis use | 1585650 | Lifestyle Survey | 3,954 | In the PAST THREE MONTHS, how often have you used Marijuana (cannabis, pot, grass, hash, weed, etc.)?<br>[Response 'Daily use'] |
|  | 434327 | Conditions | 3,712 | Cannabis abuse |
|  | 440387 | Conditions | 934 | Cannabis dependence |
|  | 440996 | Conditions | 213 | Cannabis dependence in remission |
|  | 433452 | Conditions | 143 | Cannabis dependence, continuous |
|  | 437838 | Conditions | 26 | Cannabis dependence, episodic |
|  | 4323639 | Conditions | 53 | Cannabis misuse |
|  | 4103419 | Conditions | 0 | Nondependent cannabis abuse |
|  | 435231 | Conditions | 122 | Nondependent cannabis abuse in remission |
|  | 434019 | Conditions | <20 | Nondependent cannabis abuse, continuous |
|  | 434328 | Conditions | <20 | Nondependent cannabis abuse, episodic |
| Heavy tobacco smoking | 1586162 | Lifestyle Survey | 7,373 | On average, over the entire time that you smoked, how many cigarettes did you smoke each day? (There are 20 cigarettes in a pack.)<br>[Response >20] |
| Antipsychotic prescription | 21604490 | Drugs | 31,511 | Antipsychotics |

We classified participants into different case/control groups (**Supplementary Figure 1**). First, we used “relaxed” definitions that assigned individuals to case status regardless of comorbidity (e.g., schizophrenia with or without history of heavy cannabis use). Second, to account for the comorbid phenotypic manifestation of schizophrenia and heavy cannabis use, we created “strict” definitions that excluded comorbidities (i.e., cases with heavy cannabis use histories that excluded those with a schizophrenia diagnosis; schizophrenia cases excluding those with a history of heavy cannabis use). Finally, we also defined a comorbid case group, which included those with *both* a history of heavy cannabis use *and* a schizophrenia diagnosis.

### Polygenic Analysis

All analyses were conducted using the AoU Researcher Workbench cloud computing environment. Single nucleotide polymorphisms (**SNPs**) were curated using the AoU short-read WGS Allele Count/Allele Frequency (**ACAF**) callset for each ancestral subpopulation, and further filtered to only biallelic SNPs present in HapMap3 from the European and African 1000 Genomes Linkage Disequilibrium (**LD**) Scores. Cohort-specific polygenic scores (**PGS**) were calculated using PRS-CS “auto” v1.1.0^24^ based on SNPs identified by publicly-available GWAS summary statistics of schizophrenia (AFR *N*_cases_= 7,509, *N*_controls_= 8,337^12^; EUR *N*_cases_=67,390, *N*_controls_=94,015^11^ and CUD (AFR *N*_cases_=19,065, *N*_controls_=104,143; EUR *N*_cases_=42,281, *N*_controls_=843,744^15^) and that intersected with SNPs in the AoU database. Schizophrenia and CUD PGSs were created from up to 973,634 SNPs using the allelic-scoring function, *score,* in PLINK (v1.9)^24^.

### Statistical analyses

We examined the independent and combined effect of CUD and schizophrenia PGS on both heavy cannabis use and schizophrenia case status from unrelated individuals^21^ using various case/control groups based on relaxed, strict and comorbid definitions (**Figure 1**). All models were adjusted for age, sex, and the first 10 ancestry-specific genomic PCs. Tobacco smoking is also prevalent among those with cannabis use and schizophrenia^25,26^; we performed sensitivity analyses using heavy tobacco smoking as a covariate.

**Figure 1.**
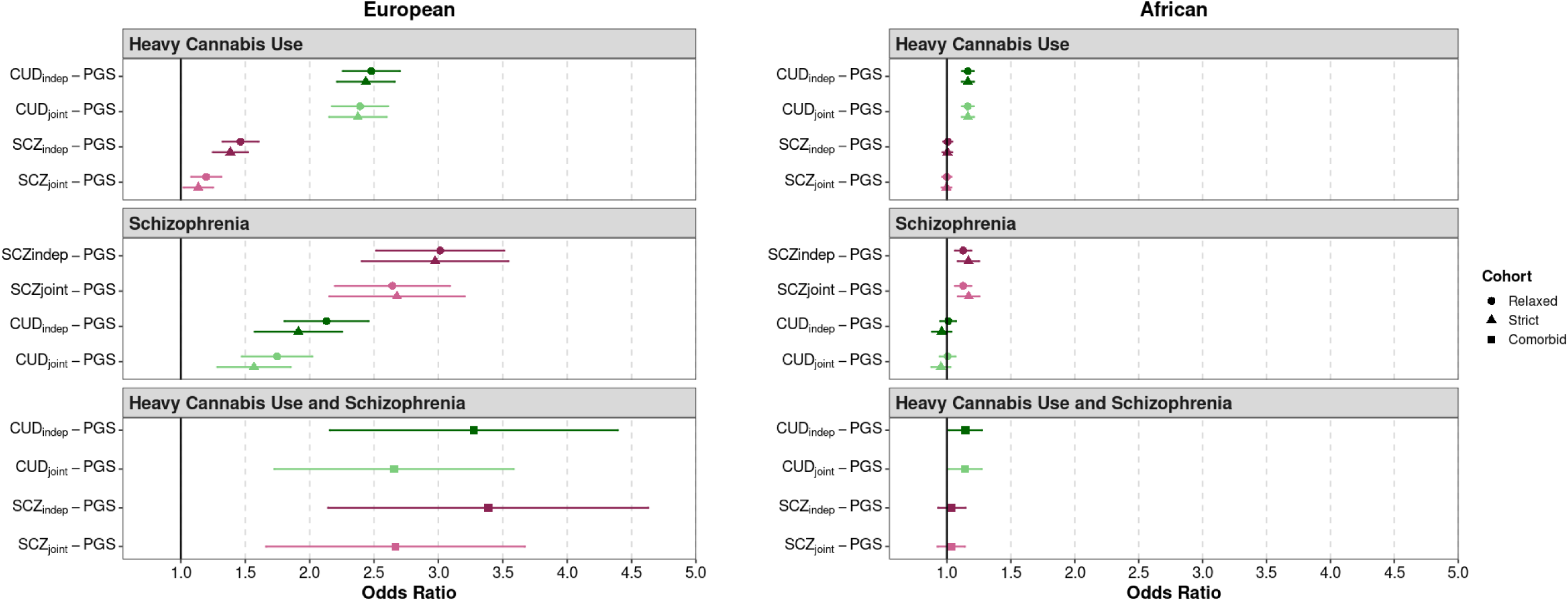
CUD and schizophrenia (SCZ) polygenic score associations with heavy cannabis use and schizophrenia. Associations shown for European and African samples under Relaxed (cases only screened for the primary outcome), Strict (cases with comorbid schizophrenia or heavy cannabis use removed), and comorbid (including both schizophrenia and heavy cannabis use) case definitions. Analyses conducted with CUD and SCZ PGSs considered independently (**indep**) or jointly (**joint**).

Liability scale *R^2^*was calculated according to Lee *et al*^27^ using the R package *fmsb* (v0.7.6), the *NagelkerkeR2* function, and the estimated population-level prevalence of 0.90% for schizophrenia^28^ and 6.27% for CUD^29^ in US adults.

## RESULTS

Participant demographics are described in **Supplementary Table 1**. For the case/control composition, see **Supplementary Figure 1**, and for the distribution of PGS across groups see **Supplementary Figures 2-3**.

### CUD-PGS and SCZ-PGS associations with heavy cannabis use

#### Relaxed case definitions

In the EUR sample, both CUD-PGS and SCZ-PGS were significantly associated with heavy cannabis use when modelled independently, with a larger magnitude of effect for CUD-PGS, as expected (CUD_ind_-PGS OR = 2.479, 95% CI: 2.267– 2.711; SCZ_ind_-PGS OR = 1.464, 95% CI: 1.33–1.611; **Table 2**, **Figure 1**). In the model including both PGS, CUD-PGS remained strongly associated with heavy cannabis use (CUD_joint_-PGS OR = 2.391, 95% CI: 2.182–2.620), with only a 3.51% reduction in the effect size when modelled alongside SCZ-PGS, while SCZ-PGS retained a significant but attenuated effect (SCZ_joint_-PGS OR = 1.198, 95% CI: 1.086–1.321), with a 18.24% reduction in the effect size compared to modelling SCZ-PGS alone.

**Table 2.**
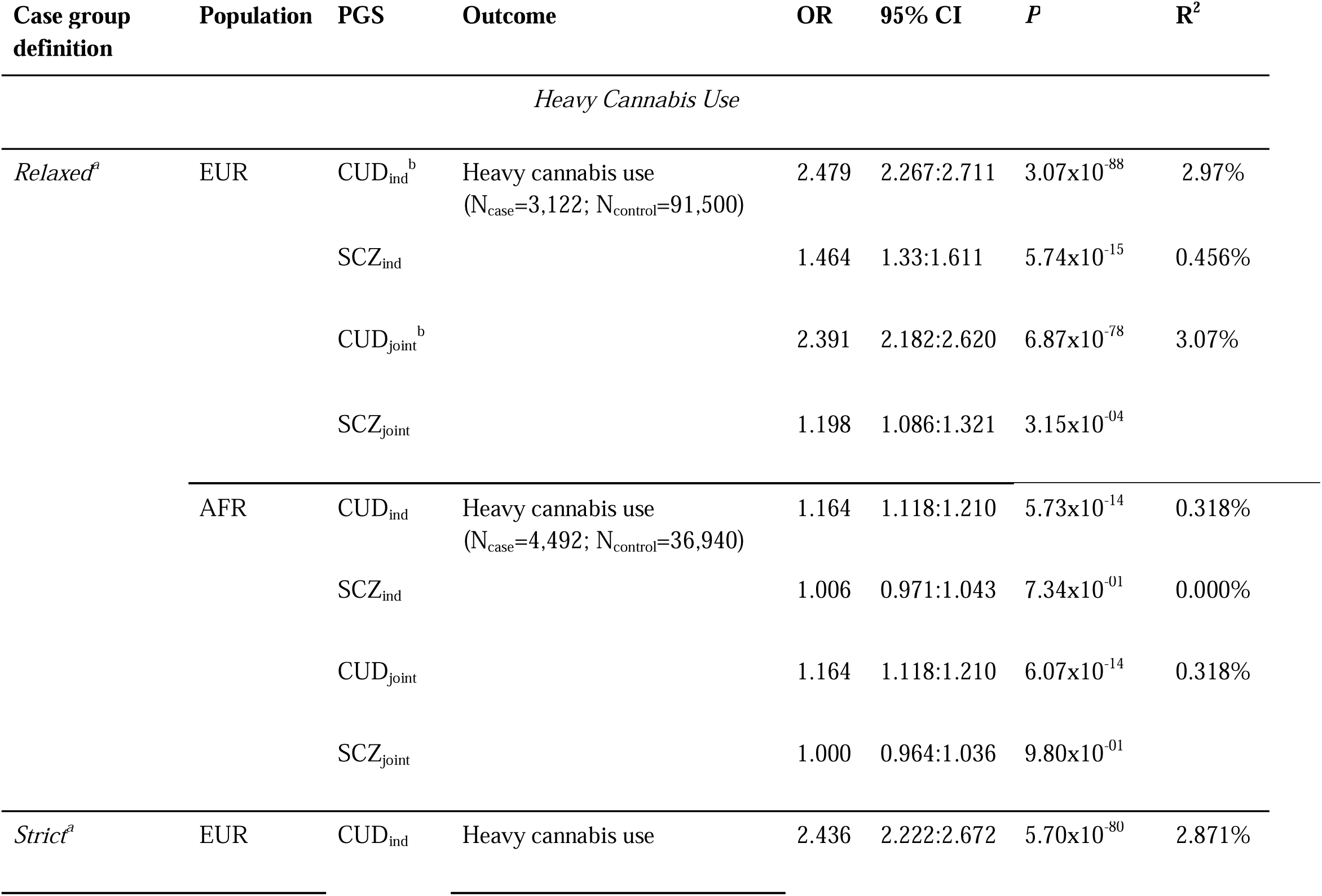
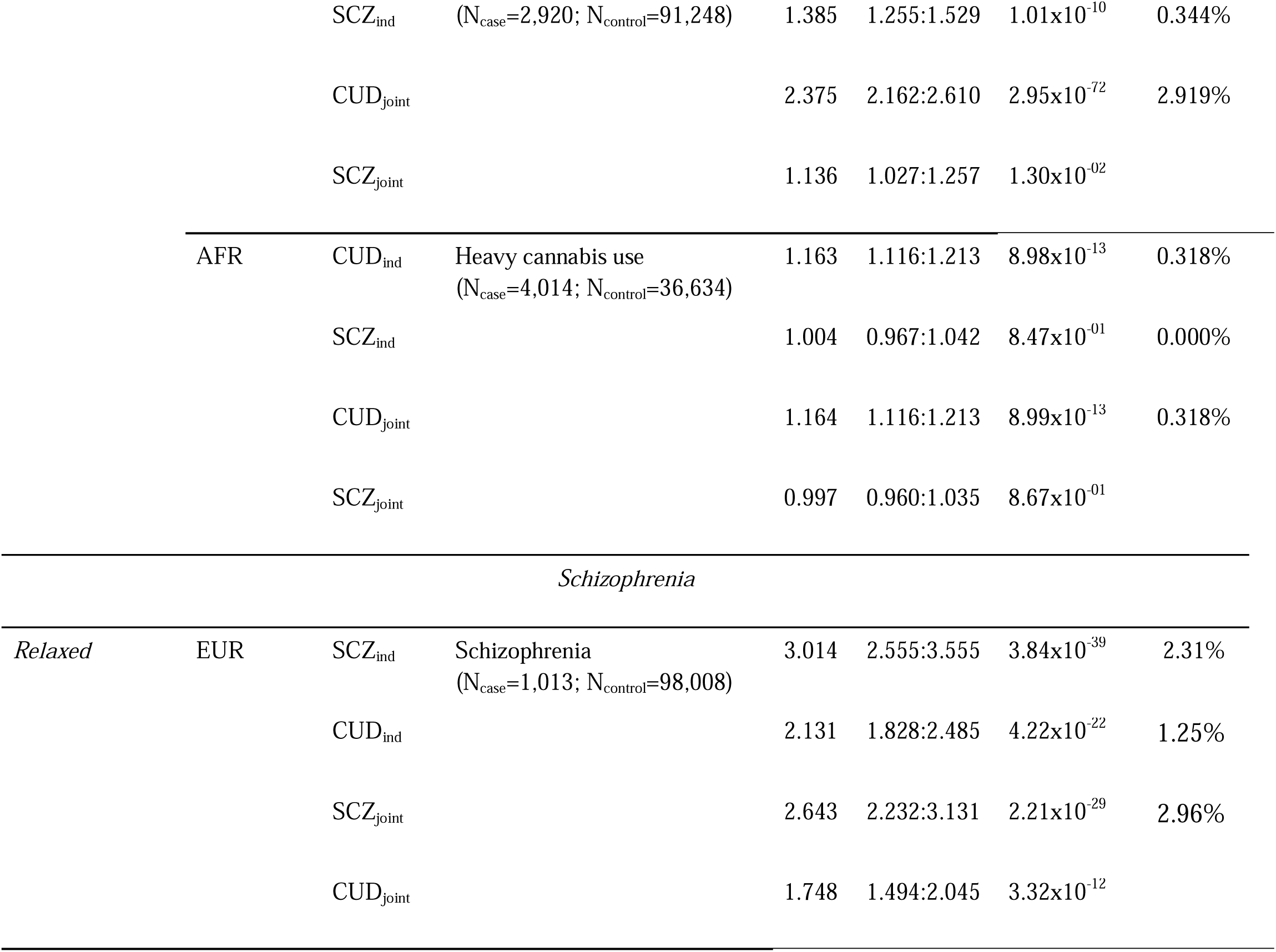
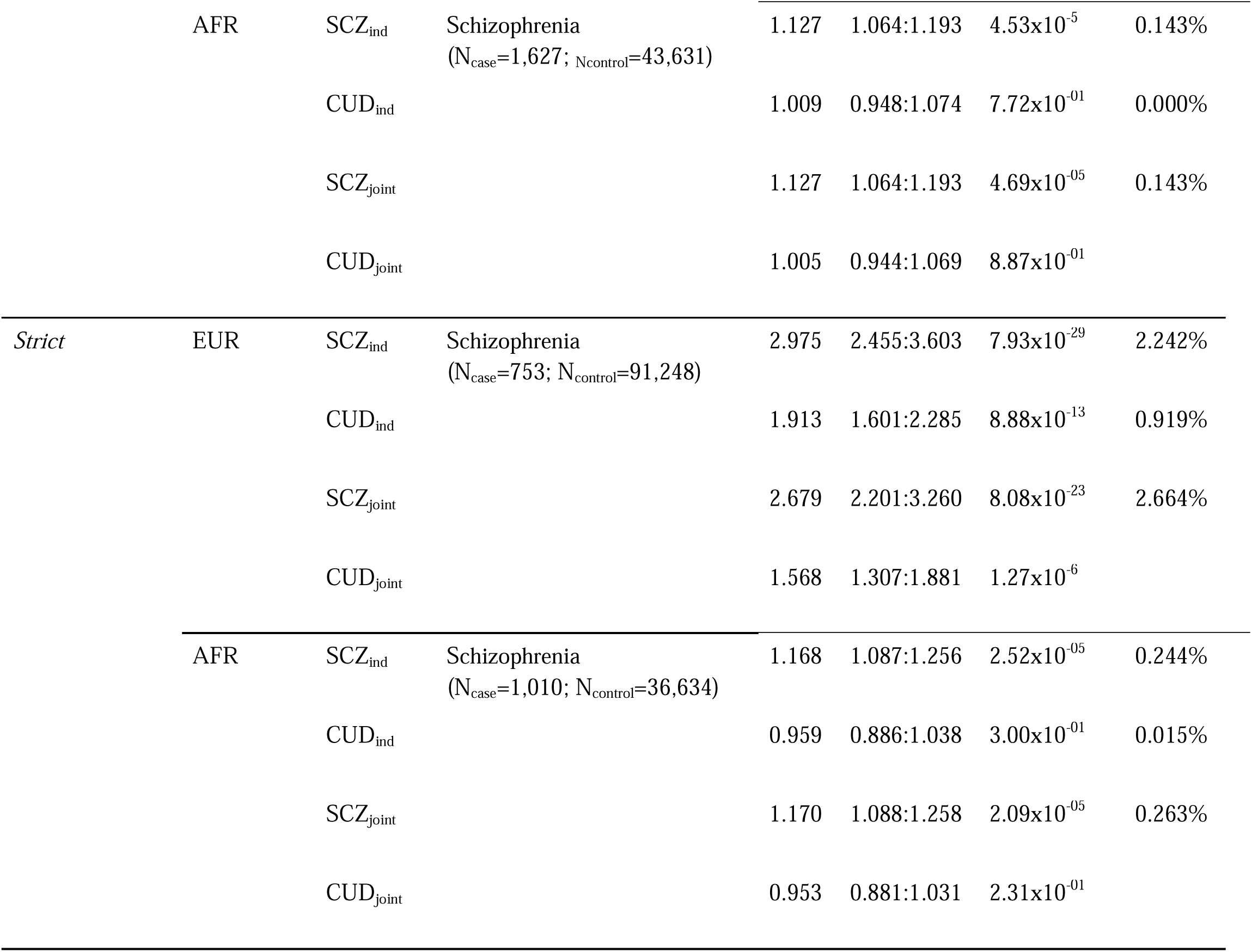
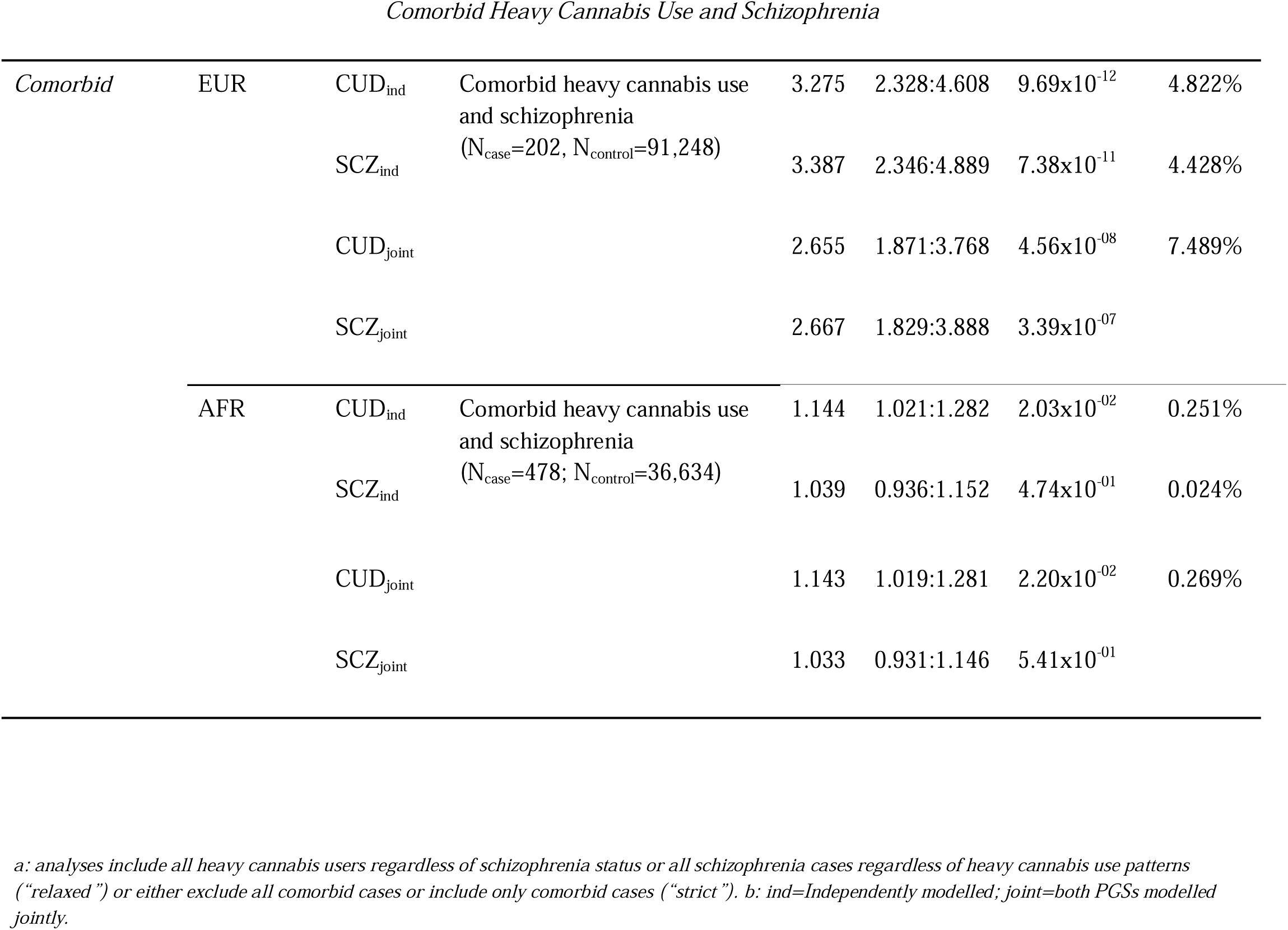
Summary of polygenic associations with heavy cannabis use and schizophrenia across various case group definitions.

In the AFR sample, CUD-PGS was also associated with heavy cannabis use both when modelled independently (CUD_ind_-PGS OR = 1.164, 95% CI: 1.118-1.210) and jointly with SCZ-PGS (CUD_joint_-PGS OR = 1.164, 95% CI: 1.118-1.210; **Table 2**, **Figure 1**). SCZ-PGS was not significantly associated with heavy cannabis use neither when modelled independently (SCZ_ind_-PGS OR = 1.006, 95% CI: 0.971–1.043) or jointly (SCZ_joint_-PGS OR = 1.000, 95% CI: 0.964–1.036).

#### Strict case definitions

We next investigated associations between CUD-PGS and SCZ-PGS and heavy cannabis use when all cases with comorbid schizophrenia were excluded. In the EUR sample, CUD-PGS remained significantly associated with heavy cannabis use (CUD_ind_-PGS OR = 2.436, 95% CI: 2.222–2.672) while SCZ-PGS showed a significant but somewhat attenuated association (SCZ_ind_-PGS OR = 1.385, 95% CI: 1.255-1.529). When both PGSs were included, CUD-PGS was associated with heavy cannabis use (CUD_joint_-PGS OR = 2.375, 95% CI: 2.162-2.610), while the SCZ-PGS association was further attenuated (OR = 1.136, 95% CI: 1.027-1.257, *p* = 0.013).

In the AFR sample, CUD-PGS was also associated with heavy cannabis use (CUD_ind_-PGS OR = 1.163, 95% CI: 1.116-1.213; **Table 2**, **Figure 1**). SCZ-PGS was not significantly associated with heavy cannabis use either when modelled independently (SCZ_ind_-PGS OR = 1.004, 95% CI: 0.967–1.042) or jointly (SCZ_joint_-PGS OR = 0.997, 95% CI: 0.960-1.035).

### CUD-PGS and SCZ-PGS associations with schizophrenia

#### Relaxed case definitions

In the EUR sample, the SCZ-PGS and CUD-PGS were significantly associated with schizophrenia when modelled independently, with a larger magnitude of effect for the within-trait associations (SCZ_ind_-PGS OR = 3.014, 95% CI: 2.555-3.555; CUD_ind_-PGS OR = 2.131, 95% CI: 1.828–2.485). The inclusion of both PGS in a joint model resulted in a slight reduction in effect size for both PGS (SCZ_joint_-PGS OR = 2.643, 95% CI: 2.232–3.131; CUD_joint_-PGS OR = 1.748, 95% CI: 1.494–2.045; **Table 2**, **Figure 1).**

In the AFR sample, the SCZ-PGS was associated with schizophrenia both when modelled independently (SCZ_ind_-PGS OR = 1.127, 95% CI: 1.064-1.193) and jointly (SCZ_joint_-PGS OR = 1.127, 95% CI: 1.064-1.193). CUD-PGS were not associated with schizophrenia neither when modelled independently (CUD_ind_-PGS OR = 1.009, 95% CI: 0.948–1.074) or jointly (CUD_joint_-PGS OR = 1.005, 95% CI: 0.944–1.069; **Table 2**, **Figure 1**).

#### Strict case definitions

Further, we considered if the explanatory power of SCZ-PGS and CUD-PGS would differ when all schizophrenia cases with a history of heavy cannabis use were excluded (**Table 2**, **Figure 1**). Among EUR participants, SCZ-PGS was highly associated with schizophrenia in both the independent and joint models (SCZ_ind_-PGS OR = 2.975, 95% CI: 2.455-3.603; SCZ_joint_-PGS OR = 2.679, 95% CI: 2.201-3.260), while CUD-PGS had a smaller but significant effect (CUD_ind_-PGS OR = 1.913, 95% CI: 1.601-2.285; CUD_joint_-PGS OR = 1.568, 95% CI: 1.307-1.881). In the AFR sample, SCZ-PGS was significantly associated with schizophrenia (SCZ_ind_-PGS OR = 1.168, 95% CI: 1.087-1.256; SCZ_joint_-PGS OR = 1.170, 95% CI: 1.088-1.258), and CUD-PGS had a weaker and non-significant effect (CUD_ind_-PGS OR = 0.959, 95% CI: 0.886–1.038; CUD_joint_-PGS OR = 0.953, 95% CI: 0.881-1.031).

### CUD-PGS and SCZ-PGS associations with comorbid heavy cannabis use and schizophrenia

Finally, we considered the explanatory value of both SCZ-PGS and CUD-PGS on cases with comorbid schizophrenia and heavy cannabis use (**Table 2**, **Figure 1**). In the EUR sample, both PGSs were associated with these comorbid cases when modelled independently (CUD_ind_-PGS OR = 3.275, 95% CI: 2.328-4.608; SCZ_ind_-PGS OR = 3.387, 95% CI: 2.346-4.889), and the effect sizes were slightly reduced for both PGS when modelled jointly (SCZ_joint_-PGS OR = 2.667, 95% CI = 1.829–3.888; CUD_joint_-PGS OR=2.655, 95% CI=1.871-3.768). In the AFR sample, we observed a similar pattern, although the SCZ-PGS was not significantly associated with these comorbid cases in either the independent or joint models (SCZ_ind_-PGS OR = 1.039, 95% CI = 0.936-1.152; SCZ_joint_-PGS OR = 1.033, 95% CI = 0.931-1.146; CUD_ind_-PGS OR = 1.144, 95% CI = 1.021-1.282; CUD_joint_-PGS OR = 1.143, 95% CI = 1.019-1.281).

## DISCUSSION

Decades of research have identified a strong association between heavy cannabis use and schizophrenia, with evidence of correlated genetic factors. However, the genetic characterization of this comorbidity is incomplete because prior studies have not fully accounted for the genetic correlation between CUD and SCZ and the phenotypic co-occurrence of these traits in target samples. In this study we modelled CUD-PGS and SCZ-PGS jointly and found that while SCZ-PGS explained little additional variance in heavy cannabis use beyond CUD-PGS (0.1% increase), CUD-PGS accounted for greater (although still modest) variance explained for schizophrenia (0.7% increase beyond the SCZ-PGS). We hypothesized more stringent case definitions would improve specificity and help delineate the extent to which underlying genetic risk contributed to CUD/schizophrenia case status. However, we showed that there was little difference in the effect sizes between the relaxed (i.e., case status regardless of comorbidity) versus strict (i.e., case status excluding comorbidity) and comorbid definitions (**Figure 1**). For instance, the association between CUD-PGS and CUD and SCZ, regardless of their comorbidity, was statistically equivalent (range 1.91 - 3.28). Even though confidence intervals were rather wide, associations between CUD and SCZ PGS were notable higher for the comorbid definition (*R^2^* up to 7.5%) compared to the relaxed and strict definitions (∼3%). These findings have important implications for schizophrenia aetiology, and demonstrate that CUD genetic liability is associated with schizophrenia risk even in individuals who do not report heavy cannabis use. Taken together, these findings suggest that prior genetic correlations between CUD and SCZ are partially due to true horizontal pleiotropy rather than confounding due to co-occurring diagnoses. In other words, regardless of comorbidity, the aetiologies of CUD and SCZ are partially due to the same genetic pathways.

When investigating the relationship between CUD-PGS and SCZ-PGS and heavy cannabis use status using traditional definitions, we identified that, as expected, within-trait PGS associations (i.e., CUD-PGS and heavy cannabis use) provided the strongest explanatory power (3% in the EUR sample; 0.3% in the AFR sample). Excluding individuals with schizophrenia case status did not increase explanatory power (2.9% in the EUR sample; 0.3% in the AFR sample). We hypothesize that this may be the case because many contributing cohorts in CUD GWAS already excluded for psychotic disorders. In contrast, SCZ-PGS showed a weak association with heavy cannabis use in the traditional analysis, and this was further reduced when CUD-PGS was included in joint models. This finding suggests that previous reports of SCZ-PGS predicting cannabis use may partially result from unmeasured comorbidity between schizophrenia risk and cannabis use^16,30,31^.

When investigating the relationship between CUD-PGS and SCZ-PGS and schizophrenia status, SCZ-PGS was associated with schizophrenia regardless of case definition (2.2-2.33% in the EUR sample; 0.5-0.6% in the AFR sample). Similarly, CUD-PGS was associated with schizophrenia regardless of case definition (0-9-1.3% in the EUR sample; 0.05% in the AFR sample). When modelled jointly, CUD-PGS explained substantial phenotypic variance of schizophrenia above and beyond that of SCZ-PGS (28.14% increase). While it is possible that individuals with higher CUD-PGS are more likely to use cannabis and that cannabis use could increase schizophrenia risk, our findings suggest that the association between CUD-PGS and schizophrenia is not solely explained by cannabis use. Instead, this reflects potential genetic liability shared between the two conditions, consistent with horizontal pleiotropy^9^. Mendelian randomization studies suggest bidirectional causal effects between CUD and schizophrenia^14,15,32^. Though this study was not designed to interrogate the causal nature of the observed associations, future analyses could build upon our work to calculate PGS that are derived from multivariate methods (i.e., genomic structural equation modelling) that parse CUD and schizophrenia genetics, or select variants based on Mendelian randomisation results^33^. These findings also caution future MR studies to carefully estimate and account for horizontal pleiotropy (e.g., MR-PRESSO^34^) when investigating the causal relationships between cannabis use and schizophrenia.

There are limitations worth noting. PGS are inherently limited by the GWAS data used to build them. While we used PRS-CSx^24^ to improve power by leveraging GWAS findings from both EUR and AFR populations (**Supplementary Table 3**), differences in predictive power of PGS in the AFR sample are likely driven by poor PGS generalisability rather than true biological or environmental effects^35^. Even among EUR it is important to note that genetic liability for these traits explains a relatively small proportion of the variance in both outcomes. Therefore, PGS are not yet clinically meaningful. Moreover, CUD is genetically correlated with a variety of other behaviours (e.g., depressed mood, executive functioning, impulsivity)^14,15^, which may co-occur with schizophrenia and thus, CUD PGS may have indexed pleiotropic risk that extends beyond associations with heavy cannabis use. Of note, while tobacco use is both correlated with CUD and schizophrenia and may be partly responsible for this residual association, adjusting for tobacco use did not alter our findings (**Supplementary Table 3**)^8^. Our reliance on EHR and/or self-report data may underestimate the true prevalence of heavy cannabis and tobacco use in our cohort (e.g., 6.7% in the general population vs ∼2% here). Lastly, the SCZ-PGS association may be due to undetected CUD in schizophrenia GWAS. The same is true for CUD GWAS, but less apparent due to the low schizophrenia prevalence. Without accounting for phenotypic overlap in the original GWASs, it will be difficult to confirm the extent to which this explains cross-trait associations.

In conclusion, we find compelling evidence of common underlying genetic mechanisms between CUD and schizophrenia. Future studies with more granular phenotyping will be essential to better understand the true extent of this putative horizontal pleiotropy.

## Data Availability

The AoU workspace used for this project will be made available upon request to registered and eligible AoU researchers through the AoU Research Workbench.

## Acknowledgements and Funding

This work was supported by the National Institute on Drug Abuse (NIDA DP1DA054394 JJM, SSR; NIDA K01DA051759 ECJ), and the California Tobacco-Related Disease Research Program (T32IR5226). JYK is supported by a Canada Research Chair in Translational Neuropsychopharmacology. AA receives funding from R01DA054869 and R21DA061592. MDF and IAZ were supported by MRC SRF (MRC MR/T007818/1).

## Conflicts of Interest

The authors declare no conflicts of interest.

## Notes

### Competing Interest Statement

The authors have declared no competing interest.

### Author Declarations

Source data was available before initiation of the study through the All of Us Research Program

## REFERENCES

1. Hasan A, von Keller R, Friemel CM, et al. Cannabis use and psychosis: a review of reviews. Eur Arch Psychiatry Clin Neurosci. 2020;270(4):403–412. doi:10.1007/s00406-019-01068-z

2. Di Forti M, Quattrone D, Freeman T. The contribution of cannabis use to variation in the incidence of psychotic disorder across Europe (EU-GEI): a multicentre case-control study. Lancet Psychiatry. 2019;6(5):427–436.

3. Di Forti M, Marconi A, Carra E. Proportion of patients in south London with first-episode psychosis attributable to use of high potency cannabis: a case-control study. Lancet Psychiatry. 2015;2(3):233–238.

4. Marconi A, Di Forti M, Lewis CM, Murray RM, Vassos E. Meta-analysis of the Association Between the Level of Cannabis Use and Risk of Psychosis. Schizophr Bull. 2016;42(5):1262–1269. doi:10.1093/schbul/sbw003

5. Large M, Sharma S, Compton MT, Slade T, Nielssen O. Cannabis use and earlier onset of psychosis: a systematic meta-analysis. Arch Gen Psychiatry. 2011;68(6):555–561. doi:10.1001/archgenpsychiatry.2011.5

6. Bagot KS, Milin R, Kaminer Y. Adolescent Initiation of Cannabis Use and Early-Onset Psychosis. Subst Abuse. 2015;36(4):524–533. doi:10.1080/08897077.2014.995332

7. Cheng W, Parker N, Karadag N, et al. The relationship between cannabis use, schizophrenia, and bipolar disorder: a genetically informed study. Lancet Psychiatry. 2023;10(6):441–451. doi:10.1016/S2215-0366(23)00143-8

8. Johnson EC, Austin-Zimmerman I, Thorpe HHA, et al. Cross-ancestry genetic investigation of schizophrenia, cannabis use disorder, and tobacco smoking. Neuropsychopharmacol Off Publ Am Coll Neuropsychopharmacol. 2024;49(11):1655–1665. doi:10.1038/s41386-024-01886-3

9. Johnson EC, Hatoum AS, Deak JD, et al. The relationship between cannabis and schizophrenia: a genetically informed perspective. Addict Abingdon Engl. 2021;116(11):3227–3234. doi:10.1111/add.15534

10. Khokhar JY, Dwiel LL, Henricks AM, Doucette WT, Green AI. The link between schizophrenia and substance use disorder: A unifying hypothesis. Schizophr Res. 2018;194:78–85. doi:10.1016/j.schres.2017.04.016

11. Trubetskoy V, Pardiñas AF, Qi T, et al. Mapping genomic loci implicates genes and synaptic biology in schizophrenia. Nature. 2022;604(7906):502–508. doi:10.1038/s41586-022-04434-5

12. Bigdeli TB, Genovese G, Georgakopoulos P, et al. Contributions of common genetic variants to risk of schizophrenia among individuals of African and Latino ancestry. Mol Psychiatry. 2020;25(10):2455–2467. doi:10.1038/s41380-019-0517-y

13. Thorpe HHA, Fontanillas P, Meredith JJ, et al. Genome-wide association studies of lifetime and frequency cannabis use in 131,895 individuals. MedRxiv Prepr Serv Health Sci. Published online June 15, 2024:2024.06.14.24308946. doi:10.1101/2024.06.14.24308946

14. Johnson EC, Demontis D, Thorgeirsson TE, et al. A large-scale genome-wide association study meta-analysis of cannabis use disorder. Lancet Psychiatry. 2020;7(12):1032–1045. doi:10.1016/S2215-0366(20)30339-4

15. Levey DF, Galimberti M, Deak JD, et al. Multi-ancestry genome-wide association study of cannabis use disorder yields insight into disease biology and public health implications. Nat Genet. Published online November 20, 2023. doi:10.1038/s41588-023-01563-z

16. Elkrief L, Lin B, Marchi M, et al. Independent contribution of polygenic risk for schizophrenia and cannabis use in predicting psychotic-like experiences in young adulthood: testing gene × environment moderation and mediation. Psychol Med. 2023;53(5):1759–1769. doi:10.1017/S0033291721003378

17. Wainberg M, Jacobs GR, di Forti M, Tripathy SJ. Cannabis, schizophrenia genetic risk, and psychotic experiences: a cross-sectional study of 109,308 participants from the UK Biobank. Transl Psychiatry. 2021;11(1):211. doi:10.1038/s41398-021-01330-w

18. Hjorthøj C, Compton W, Starzer M, et al. Association between cannabis use disorder and schizophrenia stronger in young males than in females. Psychol Med. Published online May 4, 2023:1–7. doi:10.1017/S0033291723000880

19. Abush H, Ghose S, Van Enkevort EA, et al. Associations between adolescent cannabis use and brain structure in psychosis. Psychiatry Res Neuroimaging. 2018;276:53–64. doi:10.1016/j.pscychresns.2018.03.008

20. Hjorthøj C, Posselt CM, Nordentoft M. Development Over Time of the Population-Attributable Risk Fraction for Cannabis Use Disorder in Schizophrenia in Denmark. JAMA Psychiatry. 2021;78(9):1013. doi:10.1001/jamapsychiatry.2021.1471

21. All of Us Research Program Genomics Investigators. Genomic data in the All of Us Research Program. Nature. 2024;627(8003):340–346. doi:10.1038/s41586-023-06957-x

22. All of Us Research Program Investigators, Denny JC, Rutter JL, et al. The “All of Us” Research Program. N Engl J Med. 2019;381(7):668–676. doi:10.1056/NEJMsr1809937

23. Ramirez AH, Sulieman L, Schlueter DJ, et al. The All of Us Research Program: Data quality, utility, and diversity. Patterns N Y N. 2022;3(8):100570. doi:10.1016/j.patter.2022.100570

24. Ge T, Chen CY, Ni Y, Feng YCA, Smoller JW. Polygenic prediction via Bayesian regression and continuous shrinkage priors. Nat Commun. 2019;10(1):1776. doi:10.1038/s41467-019-09718-5

25. Agrawal A, Budney AJ, Lynskey MT. The co-occurring use and misuse of cannabis and tobacco: a review. Addict Abingdon Engl. 2012;107(7):1221–1233. doi:10.1111/j.1360-0443.2012.03837.x

26. Ziedonis D, Hitsman B, Beckham JC, et al. Tobacco use and cessation in psychiatric disorders: National Institute of Mental Health report. Nicotine Tob Res Off J Soc Res Nicotine Tob. 2008;10(12):1691–1715. doi:10.1080/14622200802443569

27. Lee SH, Goddard ME, Wray NR, Visscher PM. A Better Coefficient of Determination for Genetic Profile Analysis: A Better Coefficient of Determination. Genet Epidemiol. 2012;36(3):214–224. doi:10.1002/gepi.21614

28. Perälä J, Suvisaari J, Saarni SI, et al. Lifetime prevalence of psychotic and bipolar I disorders in a general population. Arch Gen Psychiatry. 2007;64(1):19–28. doi:10.1001/archpsyc.64.1.19

29. Hasin DS, Kerridge BT, Saha TD, et al. Prevalence and Correlates of DSM-5 Cannabis Use Disorder, 2012-2013: Findings from the National Epidemiologic Survey on Alcohol and Related Conditions-III. Am J Psychiatry. 2016;173(6):588–599. doi:10.1176/appi.ajp.2015.15070907

30. Verweij KJH, Abdellaoui A, Nivard MG, et al. Short communication: Genetic association between schizophrenia and cannabis use. Drug Alcohol Depend. 2017;171:117–121. doi:10.1016/j.drugalcdep.2016.09.022

31. Hiemstra M, Nelemans SA, Branje S, et al. Genetic vulnerability to schizophrenia is associated with cannabis use patterns during adolescence. Drug Alcohol Depend. 2018;190:143–150. doi:10.1016/j.drugalcdep.2018.05.024

32. Lin BD, Pries LK, Sarac HS, et al. Nongenetic Factors Associated With Psychotic Experiences Among UK Biobank Participants: Exposome-Wide Analysis and Mendelian Randomization Analysis. JAMA Psychiatry. 2022;79(9):857–868. doi:10.1001/jamapsychiatry.2022.1655

33. Garfield V, Anderson EL. A brief comparison of polygenic risk scores and Mendelian randomisation. BMC Med Genomics. 2024;17(1):10. doi:10.1186/s12920-023-01769-4

34. Verbanck M, Chen CY, Neale B, Do R. Detection of widespread horizontal pleiotropy in causal relationships inferred from Mendelian randomization between complex traits and diseases. Nat Genet. 2018;50(5):693–698. doi:10.1038/s41588-018-0099-7

35. Atkinson EG, Bianchi SB, Ye GY, et al. Cross-ancestry genomic research: time to close the gap. Neuropsychopharmacol Off Publ Am Coll Neuropsychopharmacol. 2022;47(10):1737–1738. doi:10.1038/s41386-022-01365-7

