## Supplementary Material for "Investigating the Polygenic Relationship Between Cannabis Use and Schizophrenia in the All of Us Research Program"

Isabelle Austin-Zimmerman^1^*, Hayley HA Thorpe^2^*, John J Meredith^3^, Jibran Khokhar^2^, Tian Ge^4,5,6,7^, Marta Di Forti^1,8^, Arpana Agrawal^9^, Emma C Johnson^9#^, Sandra Sanchez-Roige^3,10,11#^

**Affiliations:**

1. Social, Genetic, and Developmental Psychiatry Centre, Institute of Psychiatry, Psychology, and Neuroscience, King’s College London, London, UK
2. Department of Anatomy and Cell Biology, Schulich School of Medicine and Dentistry, Western University, London, Ontario, Canada
3. Department of Psychiatry, University of California San Diego, La Jolla, CA, USA
4. Psychiatric and Neurodevelopmental Genetics Unit, Center for Genomic Medicine, Massachusetts General Hospital, Boston, MA
5. Center for Precision Psychiatry, Department of Psychiatry, Massachusetts General Hospital, Boston, MA
6. Department of Psychiatry, Harvard Medical School, Boston, MA
7. Stanley Center for Psychiatric Research, Broad Institute of MIT and Harvard, Cambridge, MA
8. South London and Maudsley NHS Foundation Trust, London, UK
9. Department of Psychiatry, Washington University School of Medicine, Saint Louis, MO, USA
10. Institute for Genomic Medicine, University of California San Diego, La Jolla, CA, USA
11. Department of Medicine, Division of Genetic Medicine, Vanderbilt University Medical Center, Nashville, TN, USA
    *first co-authors; #senior co-corresponding authors

### Supplementary Material

**Supplementary Figures**

**Supplementary Figure 1.** Participant numbers across case-control group compositions.

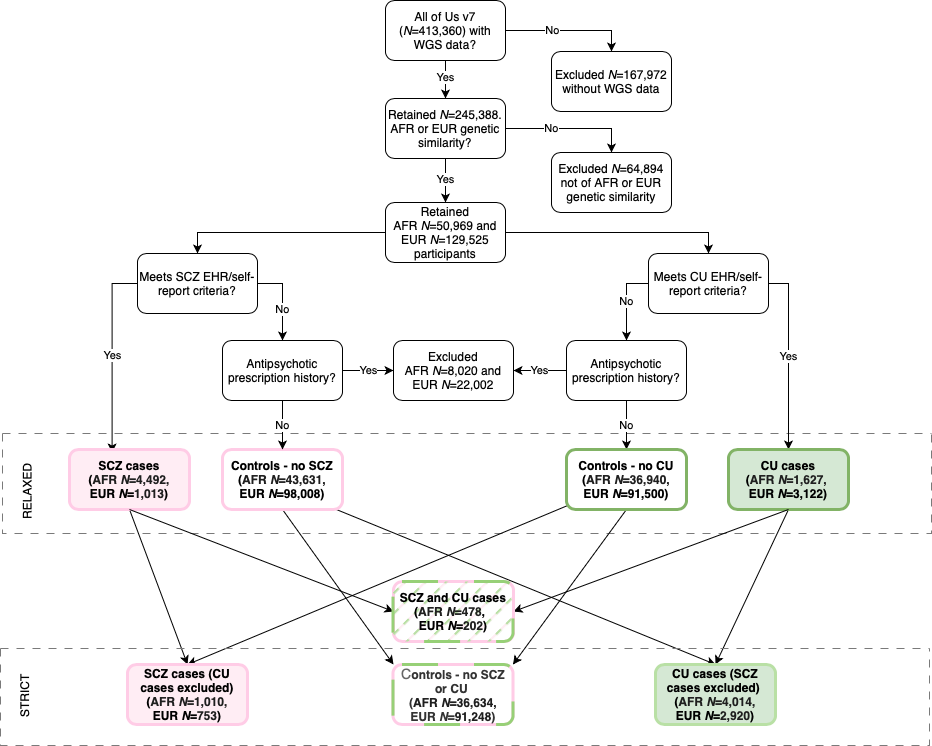

**Supplementary Figure 2.** Scaled CUD and SCZ PGSs distributions across the different case groups in the European sample.

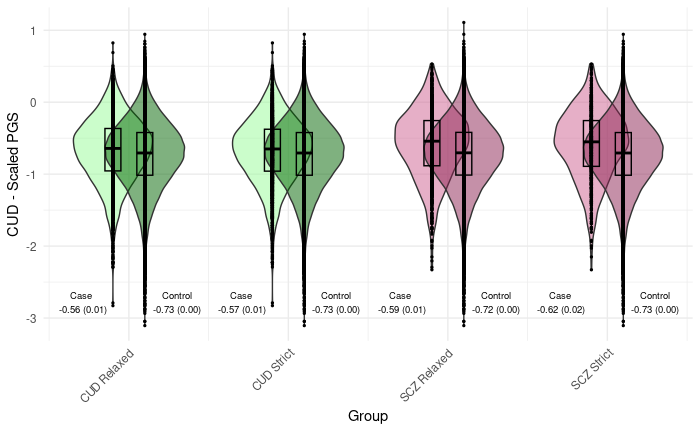

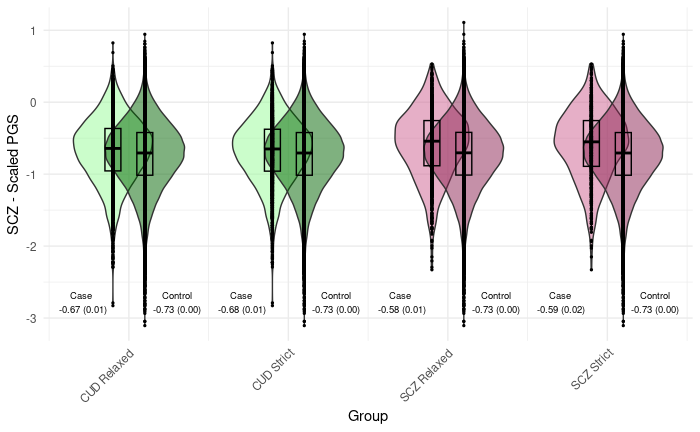

**Supplementary Figure 3.** Scaled CUD PGS distributions across the different case groups in the African sample.

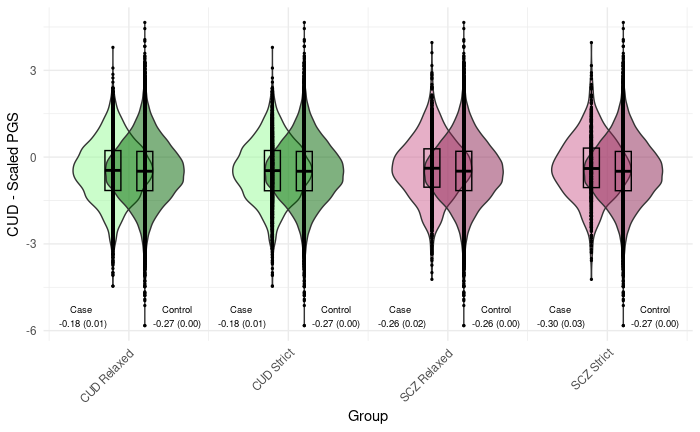

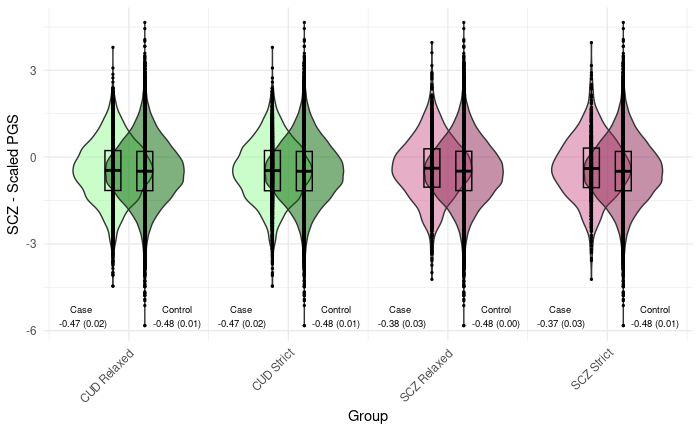
